# Estimating vaccine efficacy against transmission via effect on viral load

**DOI:** 10.1101/2021.05.03.21256556

**Authors:** Lee Kennedy-Shaffer, Rebecca Kahn, Marc Lipsitch

## Abstract

Determining policies to end the SARS-CoV-2 pandemic will require an understanding of the efficacy and effectiveness (hereafter, efficacy) of vaccines. Beyond the efficacy against severe disease and symptomatic and asymptomatic infection, understanding vaccine efficacy against transmission will help model epidemic trajectory and determine appropriate control measures. Recent studies have proposed using random virologic testing in individual randomized controlled trials to improve estimation of vaccine efficacy against infection. We propose to further use the viral load measures from these tests to estimate efficacy against transmission. This estimation requires a model of the relationship between viral load and transmissibility and assumptions about the vaccine effect on transmission and the progress of the epidemic. We describe these key assumptions, potential violations of them, and solutions that can be implemented to mitigate these violations. Assessing these assumptions and implementing this random sampling, with viral load measures, will enable better estimation of the crucial measure of vaccine efficacy against transmission.

## Main Text

To understand how vaccinations will affect the severe acute respiratory syndrome coronavirus 2 (SARS-CoV-2) pandemic, we must be able to assess both the direct protection and indirect protection that these vaccines confer (1). Individual randomized controlled trials (RCTs) conducted have shown high vaccine efficacy (VE) of several vaccine candidates in preventing symptomatic, laboratory-confirmed disease (2–5). Several studies have also demonstrated that the vaccines further protect against asymptomatic infection, as measured through virologic or serologic testing of trial participants (3–5) and through repeated sampling in longitudinal cohorts (6). However, in the absence of perfect efficacy against acquisition of the infection, fully assessing the effect of vaccination requires an assessment of the vaccine’s efficacy at preventing transmission of the pathogen from infected vaccinated individuals to susceptible individuals (7).

Throughout this paper we discuss estimands that could be estimated in studies that are randomized or observational, “idealized” or “real-world.” For brevity, we use the term efficacy throughout, to avoid the various distinctions often made between efficacy and effectiveness (8, 9, 10).

Halloran et al. described the vaccine efficacy on infectiousness and its role in understanding the total and overall vaccine effects (7). However, methods to estimate this parameter generally focus on large-scale observational studies with observation of contacts of infected individuals (8, pp. 27–28), add-on household studies, genetic linkage studies, or cluster randomized trials (1). Rinta-Kokko et al. described methods to estimate vaccine efficacy on carriage prevalence through measured odds ratios (11). More recently, similar methods have been proposed to estimate the vaccine efficacy on prevalent infection, accounting for vaccine effects that change the duration of infection (and thus, likely, the duration of infectiousness) by incorporating random sampling and testing among RCT participants (12, 13). Follman and Fay moreover describe how, by measuring viral loads or a proxy thereof, the vaccine efficacy on transmission could be estimated (13).

We describe a method, similar to that laid out in (13), to estimate vaccine efficacy against transmission by estimating both vaccine efficacy against detectable infection and vaccine efficacy against per-contact infectiousness. This process relies on sampling and virologic testing of the full study population or a random sample thereof within a RCT of a vaccine candidate, as was done in at least one vaccine study for COVID-19 (3). While Follman and Fay focus on the inferential method and statistical properties of such an estimator (13), we focus here on assumptions about the infection and transmission process and the vaccine effect that are necessary for consistent estimation of this effect.

In the next section, we describe the formulation of the estimand of interest and its relation to other common vaccine efficacy measures. In the following section, we define an estimator of this effect and show that it is consistent for that estimand. In the “Assumptions for Consistency” section, we detail the assumptions needed for that consistency, describing their implications, examples of possible violations, and approaches that may be used to mitigate these violations. Finally, we describe the implications of this method and the research needed to best understand when these assumptions are met for SARS-CoV-2 and other infectious pathogens.

## Estimand of Interest

We are interested in the vaccine efficacy in preventing transmission, *VE*_*T*_, the reduction in the probability of transmission at any given time caused by vaccination. This probability, at any time *t*, can be decomposed as follows:

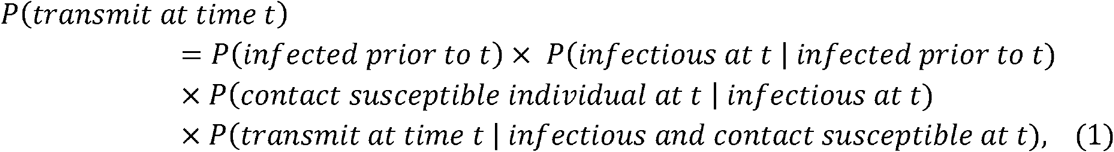

and so:

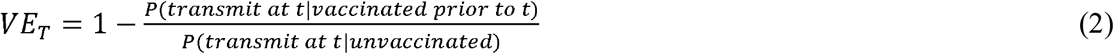

Under the assumptions that: vaccination status does not affect the probability of having contact with a susceptible individual given that an individual is infectious (Assumption 1); and that the incidence rate is constant across time (Assumption 2), the vaccine efficacy against transmission is given by:

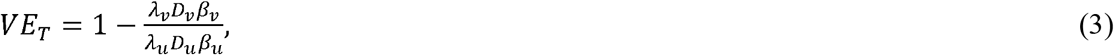

where *λ*_*ν*_ is the incidence rate of infection among vaccinated individuals, *D*_*ν*_ is the average duration of infectiousness for infected vaccinated individuals, *β*_*ν*_ is the average per-contact transmission probability from an infected vaccinated individual, and these quantities with subscript *u* refer to the quantities among unvaccinated individuals.

Mathematically, this can be rewritten as:

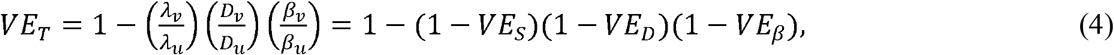

where 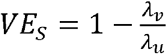 is the vaccine efficacy against susceptibility or viral acquisition, 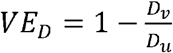 is the vaccine efficacy against duration of infectiousness, and 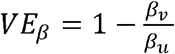 is the vaccine efficacy against per-contact infectiousness, defined as the reduction in the probability of transmission per contact (7, 8, pp. 19–29). Halloran et al. use *VE*_*I,p*_ where we use *VE*_*β*_; we avoid that notation to avoid confusion with vaccine efficacy against progression (7, 8, p. 23).

Under the assumption that an individual is infectious only if they have detectable viral load (Assumption 3), then we can write:

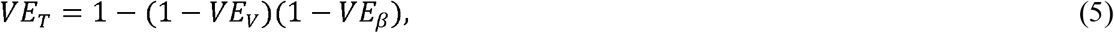

where *VE*_*V*_ =1 − (1 − *VE*_*s*_) (1 − *VE*_*D*_) is the vaccine efficacy against prevalent detectable infection, per Lipsitch and Kahn (12), and *VE*_*β*_ is the vaccine efficacy against per-contact infectiousness, as defined above.

### Estimator Using Random Samples from Vaccine RCT

By equation 5, to estimate *VE*_*T*_, it suffices to estimate *VE*_*V*_ and *VE*_*β*_. In a vaccine RCT, *VE*_*V*_ can be estimated using the odds ratio of prevalent detectable infection at time *t* among vaccinated individuals vs. unvaccinated individuals (11–13).

If a random sample of participants have virologic tests conducted at some time *t* after enrollment in a RCT, then the measured viral loads from these test results can be used to estimate the distribution of true viral loads among the vaccinated and the unvaccinated individuals. As above, we assume that Assumptions 1–3 hold. If we also assume that the per-contact infectiousness of an individual among vaccinated individuals is some known function *g*_*ν*_ (*y*) of the measured viral load *y*, and the per-contact infectiousness of an individual among unvaccinated individuals is some known function *g*_*u*_ (*y*) (Assumption 4), then:

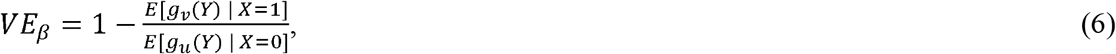

where *Y* is the measured viral load of a randomly chosen infectious individual and *X* is the vaccination status of that individual, where *X* = 1 denotes a vaccinated individual and *X* = 0 an unvaccinated individual. Note that if vaccination only affects infectiousness through its effect on viral load (that is, viral load fully mediates the vaccine efficacy on infectiousness), then *g*_*ν*_ = *g*_*u*_. These assumptions are described in more detail below.

Define the estimator:

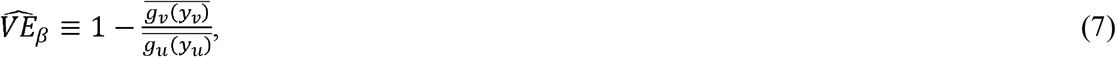

where *y*_*ν*_ is the measured viral load of an individual in the vaccinated arm of the trial and *y*_*u*_ the measured viral load of an individual in the unvaccinated arm of the trial, and the averages are across all sampled individuals in the respective arms of the trial. Then if the infectiousness functions (*g*_*v*_ and *g*_*u*_) are correctly specified with respect to the measured viral load in the sample, 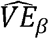 is a consistent estimator of *VE*_*β*_.

If a measured covariate (e.g., symptom status) modifies the effect of viral load on per-contact infectiousness, this can be incorporated by specifying infectiousness functions for each level of the covariate. For example, let *s* denote the symptom status of an individual (equal to 1 if symptomatic and 0 if not). Then, if the infectiousness function among symptomatic individuals, whether vaccinated or not, is given by *g*_*1*_ and the infectiousness function among individuals not currently symptomatic is given by *g*_*0*_, then:

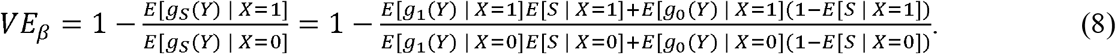

This can be estimated by:

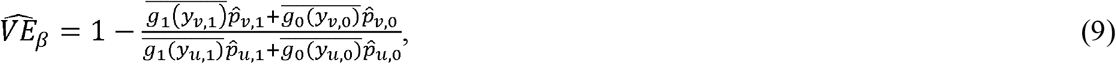

where *yi,j* represents the measured viral load of an individual in arm *i* with symptom status *j*, the overbars represent averages over the sampled individuals in that arm with that symptom status, and 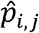 is the proportion of individuals in arm *i* who have symptom status *j*, for *i* ∈ {*u ν*} and *j* ∈, {*0*,1}.

If a study is conducted with randomly-sampled virologic tests conducted at a time point *t*, the conditions for a consistent estimator 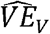 are met (12), and the conditions described above for a consistent estimator 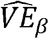 are met, then these can be combined to get a consistent estimator of *VE*_*T*_:

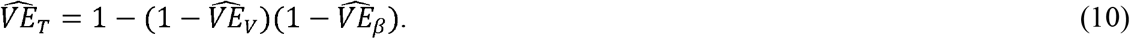

### Assumptions for Consistency

As derived in the previous section, the consistency of 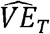 as an estimator of *VE*_*T*_ relies on four key assumptions. In this section, we describe these assumptions, the consequences of violations of the assumptions, examples of potential violations, and solutions to address these violations. Table 1 provides a summary of these assumptions.

**Table 1.**
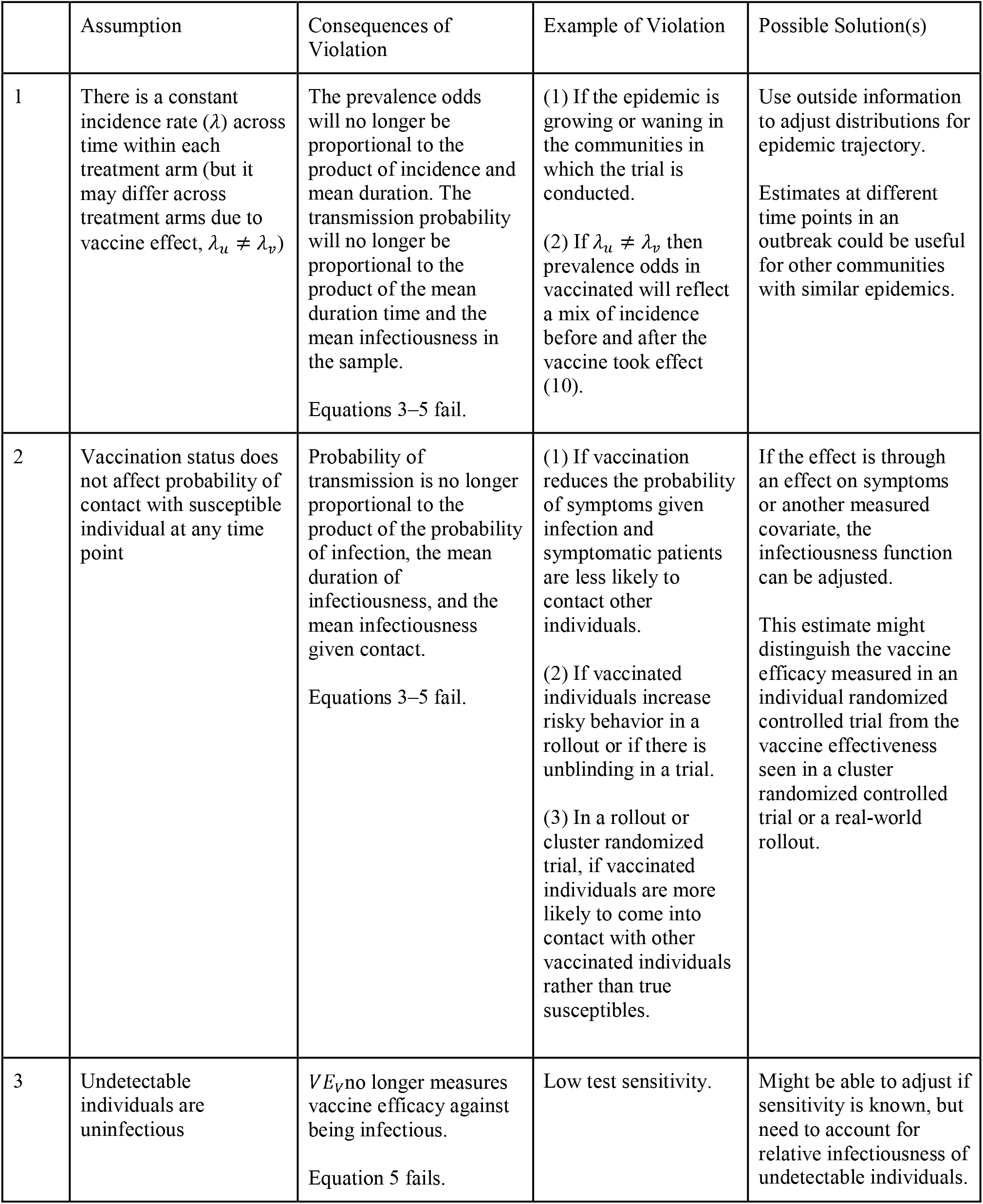
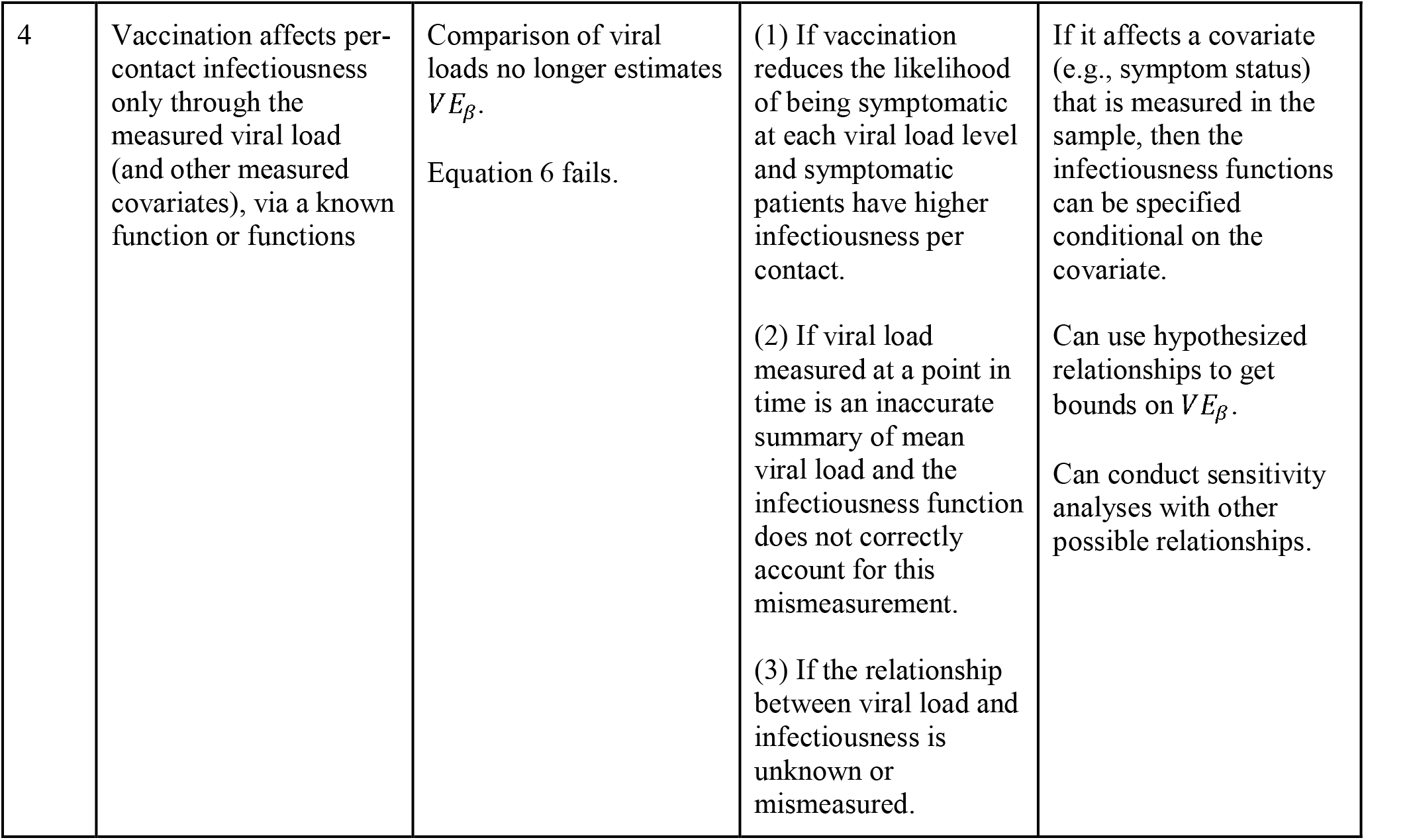
Summary of Assumptions for Consistent Estimation.

#### Assumption 1: Within each arm of the trial, incidence rate among participants is constant throughout time

Under this assumption, there is a baseline incidence rate within each arm of the vaccine trial, which differs between the arms only due to the vaccine efficacy against susceptibility. If this assumption is violated, then the distribution of viral loads in each arm, *Yν* and *Y*_*u*_, measured by cross-sectional testing at a single point in time are not representative of the distribution of viral loads across the course of infection (14).

This assumption is violated if the epidemic is growing or waning in the communities in which the trial is conducted. For example, in a growing epidemic, the average time since infection for cases ascertained in a cross-sectional sample is lower than in a flat epidemic (14–16). If the vaccine efficacy on viral load mostly affects the later stage of infection (e.g., by allowing faster viral clearance), *VE*_*β*_ will be underestimated in this sample. In a waning epidemic, more individuals will be in the later stage of infection, and so *VE*_*β*_ will be overestimated in this sample. If, on the other hand, the vaccine efficacy on viral load mostly affects the earlier stage of infection (e.g., by reducing the peak viral load), the reverse will happen: *VE*_*β*_ will be underestimated in a growing epidemic and overestimated in a waning epidemic. Data on epidemic trajectory could be used to adjust for this bias. Follman and Fay consider this bias for some viral load functions and vaccine efficacy parameters and find that the bias is less than 10% (13).

Without adjusting for this bias, the estimate represents the instantaneous vaccine effectiveness on transmission at the time of sampling. That is, the estimand is a function of the epidemic conditions in the community where the trial was conducted. This quantity might be useful in generalizability to communities at a similar stage of their epidemic trajectory.

This assumption is complicated if there are multiple variants of the pathogen with different incidence rates in the study population. In particular, even if the overall incidence of infection is constant, the assumptions can be violated if one variant is increasing while another is decreasing. If the vaccine has different efficacy levels against the variants, as has been suggested for several SARS-CoV-2 vaccines (17–19), the observed vaccine efficacy represents a weighted average of these efficacies. However, if the variants have different viral load patterns in infection, as has been suggested for multiple SARS-CoV-2 variants (20–22), the observed efficacy would then be a weighted average of the specific point-in-time efficacies against each variant at its current growth or decline rate of incidence, which is unlikely to be generalizable to any other population.

#### Assumption 2: vaccination status does not affect the probability of contact with a susceptible individual at any point in time

Under this assumption, individuals in both arms have the same distribution of contact rates. If this assumption is violated, then the probability of transmission is no longer proportional to the product of the probability of infection, the mean duration of infectiousness, and the mean per-contact infectiousness (i.e., equations 3 and 4 no longer hold). Correcting for violations of this assumption is challenging as the effects depend on changes in behavior as a result of vaccination.

For example, this violation may occur, even in a RCT with concealed allocation, if vaccination reduces the probability of symptoms given infection (VE for progression to symptoms > 0) and symptomatic individuals are less likely to contact others than asymptomatic individuals. In this case, data on symptom status of cases could be used to adjust for this effect. Additionally, risk compensation of vaccinated individuals, either during vaccine rollout or in an open-label trial (or a trial with concealed allocation but strong reactogenicity in the vaccine arm), may also affect contact patterns, violating this assumption.

During vaccine rollout, vaccinated individuals may also have different baseline behaviors from those who are not vaccinated, biasing observational estimates of vaccine effectiveness. Vaccinated individuals may be more likely to interact with other vaccinated individuals, either intentionally or by nature of social networks (e.g., in a cluster randomized trial or during geographically phased vaccination rollout). Contact surveys or proximity data from mobile phones could be used to understand the magnitude of this effect. These potential changes in behavior and contacts may differentiate the vaccine efficacy measured in an individual RCT from the vaccine effectiveness during a wide rollout or cluster randomized trial.

This assumption can be slightly relaxed if the infectiousness function relating measured viral load and per-contact infectiousness actually measures infectiousness accounting for changes in contact patterns that are fully mediated by changes in measured viral load. For example, suppose that higher viral loads are associated with more severe illness, limiting contacts per time infected. If the function *g* was estimated from a study that estimated secondary attack rate as a function of viral load (23), this change in contact pattern, mediated by the viral load, would be incorporated into *g* and thus into the vaccine efficacy estimate here. Interpretation of 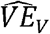 should consider the appropriate interpretation of *g*.

#### Assumption 3: individuals without detectable viral load are not infectious

This assumption implies a perfectly sensitive test for infectiousness. Under a violation of this assumption, for example because of low sensitivity, 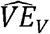 will be biased (12). Adjustments for test characteristics could alleviate the inconsistency in 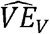. However, the estimate of 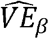 must also be adjusted to account for the infectiousness of undetected infections. This adjustment is less straightforward, as it requires an estimate for the infectiousness of individuals who are infectious but have an undetectable viral load. Sensitivity analysis could be conducted with different assumptions about the infectiousness of these individuals, e.g., by assuming they are no more infectious than the average individual with a detectable viral load.

Note that this does not make any assumption about the specificity of the test used. Imperfect specificity can cause bias in 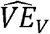, but if this is properly accounted for in the infectiousness function, as described under the next assumption, the estimator 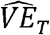 will remain consistent.

#### Assumption 4: vaccination affects per-contact infectiousness only through the measured viral load (and other measured covariates); this relationship follows a specified function or functions

Correct specification of the relationship between the measured viral load and infectiousness is crucial to the estimation of *VE*_*β*_. There are several reasons this assumption may be violated.

First, this assumption would be violated if vaccination reduces the likelihood of being symptomatic at each viral load level and symptomatic patients have higher infectiousness per contact. Data on symptom status could be used to adjust for this and perhaps separate models of the relationship could be used based on symptom status, as described above. This requires, however, the specification of more infectiousness functions.

Second, this assumption would be violated if measured viral load is not an accurate summary of the true viral load and the infectiousness function does not capture this measurement error. This could occur, for example, if the relationship between viral load and infectiousness is estimated using a specific testing platform and the viral loads are measured with a different platform that has different measurement error properties. It could also occur if the relationship is estimated from viral load measurements at a single time point during infection (e.g., symptom onset), as that will be an imperfect representation of the full trajectory of viral loads during infection.

More generally, the relationship between infectiousness and viral load may be misspecified or unknown. This could occur because the relationship is specified on the wrong scale (e.g., proportional to the log viral load or Ct value rather than to the linear viral load, or a threshold effect), because of measurement error (e.g., the discrete nature of Ct values), or uncertainty in the relationship. It might still be possible, however, to get a range of reasonable estimates for *VE*_*β*_ by conducting sensitivity analyses with a variety of potential infectiousness functions. 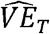 can then be bounded by calculating its value with extreme possibilities of these 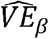 values. For example, infectiousness proportional to the viral load and infectiousness constant above a relatively low viral load threshold may be reasonable bounds for most pathogens. These bounds would, for example, encompass a true infectiousness function that is proportional to the log of the viral load.

Violations can occur specifically if vaccination itself affects the relationship between viral load and infectiousness in a way that is not mediated by symptom status or other measured covariates. Often, the only available data to estimate the infectiousness function will be from studies conducted prior to vaccination, so there may be no ability to account for this difference. For example, Regev-Yochay et al. find that the PCR-positive individuals with a Ct value below 30 were significantly less likely to test positive for antigen if vaccinated than if unvaccinated (24). If antigen tests are a more accurate reflection of infectiousness than PCR positivity with Ct values below 30, then this would indicate that vaccines can reduce transmissibility through a mechanism not mediated fully by PCR viral load.

In practice, this element of the analysis will rely on epidemiological studies conducted to estimate this relationship (23, 25), and laboratory analyses that improve biological understanding of the transmission process (26). There will always be uncertainty in the measurement of this relationship. Accounting for this uncertainty can occur via the sensitivity analyses described above or by formal Bayesian inference that incorporates prior distributions on the infectiousness function.

## Discussion

Understanding the efficacy of vaccines and how they will affect the trajectory of the SARS-CoV-2 pandemic requires a wide array of studies, both randomized and observational. To get the most information out of large-scale RCTs, these trials should incorporate virologic testing of a random sample of participants, as suggested previously (12, 13). This allows estimation of vaccine efficacy against prevalent infection, rather than just symptomatic infection. We have shown that, under additional assumptions, using the quantitative information from such random testing additionally allows for the estimation of vaccine efficacy against transmission, a crucial parameter for understanding how widespread vaccination will affect circulating virus.

This estimation requires four key assumptions. First, the incidence rate among participants must be constant throughout time within each arm of the trial. Second, vaccination status must not affect the probability of contact with a susceptible individual during the study. Third, individuals without a detectable viral load are assumed to be uninfectious. And fourth, vaccination must affect infectiousness only through a known function of the measured viral load and, potentially, other measured covariates.

Meeting the assumptions for this estimation requires an understanding of the role of viral load in the transmission process. This increased understanding of viral load and proxies such as the cycle threshold measured by polymerase chain reaction tests can contribute to better understanding of the pandemic in several ways, including monitoring population incidence (14), assessing the role of variants in outbreaks (20, 22), assessing test performance (27), and clinical management (28). While some studies have assessed the association between measured viral load and infectiousness (23, 25), more such work is needed to improve this aspect of the estimation.

This method could be used to compare the efficacy of two different vaccines or vaccine regimens and to observational studies, in which control of confounding and avoidance of selection biases will present challenges similar to, but slightly more extensive than, the ones in standard vaccine effectiveness studies (29). Such an analysis would require exchangeability assumptions as well as the assumptions described above: that there is no unmeasured confounding between vaccination status and any of the following: the probability of infection, duration of infection, measured viral load, and the relationship between viral load and infectiousness. Moreover, for a retrospective study, selection criteria would need to not exclude individuals because of any factor—e.g., hospitalization or death—causally related to vaccination and measured viral load (30).

Ending the SARS-CoV-2 pandemic and mitigating future pandemics will require the best available evidence, as quickly as possible, on the full set of measures of vaccine efficacy. We have described a method to gain a richer picture of vaccine efficacy on transmission with the optimal use of quantitative data from virologic testing on a sample of trial participants.

## Data Availability

No data referred to in manuscript.

## References

[1] Lipsitch M and Dean NE. Understanding COVID-19 vaccine efficacy. Science 2020; 370: 763–765.

[2] Polack FP, Thomas SJ, Kitchin N, et al. Safety and efficacy of the BNT162b2 mRNA Covid-19 vaccine. N Engl J Med 2020; 383: 2603–2615.

[3] Voysey M, Clemens SAC, Madhi SA, et al. Safety and efficacy of the ChAdOx1 nCoV-19 vaccine (AZD1222) against SARS-CoV-2: an interim analysis of four randomised controlled trials in Brazil, South Africa, and the UK. Lancet 2021; 397: 99–111.

[4] Baden LR, El Sahly HM, Essink B, et al. Efficacy and safety of the mRNA-1273 SARS-CoV-2 vaccine. N Engl J Med 2021; 384: 403–416.

[5] Oliver SE, Gargano JW, Scobie H, et al. The Advisory Committee on Immunization Practices’ interim recommendation for use of Janssen COVID-19 vaccine—United States, February 2021. MMWR Morb Mortal Wkly Rep 2021; 70: 329–332.

[6] Thompson MG, Burgess JL, Naleway AL, et al. Interim estimates of vaccine effectiveness of BNT162b2 and mRNA-1273 COVID-19 vaccines in preventing SARS-CoV-2 infection among health care personnel, first responders, and other essential and frontline workers—eight U.S. locations, December 2020–March 2021. MMWR Morb Mortal Wkly Rep 2021; 70: 495–500.

[7] Halloran ME, Longini IM, and Struchiner CJ. Design and interpretation of vaccine field studies. Epidemiol Rev 1999; 21: 73–88.

[8] Halloran ME, Longini IM, and Struchiner CJ. Design and Analysis of Vaccine Studies. 2010. New York: Springer.

[9] Gartlehner G, Hansen RA, Nissman D, Lohr KN, Carey TS. Criteria for Distinguishing Effectiveness from Efficacy Trials in Systematic Reviews. AHRQ Publication No. 06-0046. 2006. Rockville, MD: Agency for Healthcare Research and Quality.

[10] Singal AG, Higgins PDR, Waljee AK. A primer on effectiveness and efficacy trials. Clin Transl Gastroenterol 2014; 5: e45.

[11] Rinta-Kokko H, Dagan R, Givon-Lavi N, and Auranen K. Estimation of vaccine efficacy against acquisition of pneumococcal carriage. Vaccine 2009; 27: 3831–3837.

[12] Lipsitch M and Kahn R. Interpreting vaccine efficacy trial results for infection and transmission. medRxiv Preprint 2021; DOI:10.1101/2021.02.25.21252415.

[13] Follman D and Fay M. Vaccine efficacy at a point in time. medRxiv Preprint 2021; DOI:10.1101/2021.02.04.21251133.

[14] Hay JA, Kennedy-Shaffer L, Kanjilal S, et al. Estimating epidemiologic dynamics from cross-sectional viral load distributions. medRxiv Preprint 2021; DOI:10.1101/2020.10.08.20204222.

[15] Wallinga J, Lipsitch M. How generation intervals shape the relationship between growth rates and reproductive numbers. Proc R Soc B 2007; 274: 599–604.

[16] Rydevik G, Innocent GT, Marion G, et al. Using combined diagnostic test results to hindcast trends of infection from cross-sectional data. PLoS Comput Biol 2016; 12: e1004901.

[17] Madhi SA, Baillie V, Cutland CL, et al. Efficacy of the ChAdOx1 nCoV-19 Covid-19 vaccine against the B.1.351 variant. N Engl J Med 2021; DOI:10.1056/NEJMoa2102214.

[18] Mahase E. Covid-19: Novavax vaccine efficacy is 86% against UK variant and 60% against South African variant. BMJ 2021; 372: 296.

[19] Planas D, Bruel T, Grzelak L, et al. Sensitivity of infectious SARS-CoV-2 B.1.1.7 and B.1.351 variants to neutralizing antibodies. Nat Med 2021; DOI:10.1038/s41591-021-01318-5.

[20] Kidd M, Richter A, Best A, et al. S-variant SARS-CoV-2 lineage B1.1.7 is associated with significantly higher viral loads in samples tested by ThermoFisher TaqPath RT-qPCR. J Infect Dis 2021; DOI:10.1093/infdis/jiab082.

[21] Kissler SM, Fauver JR, Mack C, et al. Densely sampled viral trajectories suggest longer duration of acute infection with B.1.1.7 variant relative to non-B.1.1.7 SARS-CoV-2. medRxiv Preprint 2021; DOI:10.1101/2021.02.16.21251535.

[22] Faria NR, Mellan TA, Whittaker C, et al. Genomics and epidemiology of the P.1 SARS-CoV-2 lineage in Manaus, Brazil. Science 2021; DOI:10.1126/science.abh2644.

[23] Lee LYW, Rozmanowski S, Pang M, et al. SARS-CoV-2 infectivity by viral load, S gene variants and demographic factors and the utility of lateral flow deviices to prevent transmission. medRxiv Preprint 2021; DOI:10.1101/2021.03.31.21254687.

[24] Regev-Yochay G, Amit S, Bergwerk M, et al. Decreased infectivity following BNT162b2 vaccination. SSRN Preprint 2021; DOI:10.2139/ssrn.3815668.

[25] Mitjà O, Corbacho-Monné M, Ubals M, et al. A cluster-randomized trial of hydroxychloroquine for prevention of Covid-19. N Engl J Med 2021; 384: 417–427.

[26] Singanayagam A, Patel M, Charlett A, et al. Duration of infectiousness and correlation with RT-PCR cycle threshold values in cases of COVID-19, England, January to May 2020. Euro Surveill 2020; 25: 2001483.

[27] Tom MR, Mina MJ. To interpret the SARS-CoV-2 test, consider the cycle threshold value. Clin Infect Dis 2020; 71: 2252–2254.

[28] Moraz M, Jacot D, Papadimitriou-Olivgeris M, et al. Universal admission screening strategy for COVID-19 highlighted the clinical importance of reporting SARS-CoV-2 viral loads. New Microbes New Infect 2020; 38: 100820.

[29] Lipsitch M, Jha A, Simonsen L. Observational studies and the difficult quest for causality: lessons from vaccine effectiveness and impact studies. Int J Epidemiol 2016; 45: 2060–2074.

[30] Accorsi EK, Qiu X, Rumpler E, et al. How to detect and reduce potential sources of biases in studies of SARS-CoV-2 and COVID-19. Eur J Epidemiol 2021; 36: 179–196.

